# Variance-quantitative trait loci enable systematic discovery of gene-environment interactions for cardiometabolic serum biomarkers

**DOI:** 10.1101/2021.11.08.21265930

**Authors:** Kenneth E. Westerman, Timothy D. Majarian, Franco Giulianini, Dong-Keun Jang, Jose C. Florez, Han Chen, Daniel I. Chasman, Miriam S. Udler, Alisa K. Manning, Joanne B. Cole

## Abstract

Gene-environment interactions (GEIs) represent the modification of genetic effects by environmental exposures and are critical for understanding disease and informing personalized medicine. GEIs often induce differential phenotypic variance across genotypes; these variance-quantitative trait loci (vQTLs) can be prioritized in a two-stage GEI detection strategy to greatly reduce the computational and statistical burden and enable testing of a broader range of exposures. We performed genome-wide vQTL analysis for 20 serum cardiometabolic biomarkers by multi-ancestry meta-analysis of 350,016 unrelated participants in the UK Biobank, identifying 182 independent locus-biomarker pairs (*p* < 4.5×10^−9^). Most vQTLs were concentrated in a small subset (4%) of loci with genome-wide significant main effects, and 44% replicated (*p* < 0.05) in the Women’s Genome Health Study (N = 23,294). Next, we tested each vQTL for interaction across 2,380 exposures, identifying 846 significant GEIs (*p* < 2.4×10^−7^). Specific examples demonstrated interaction of triglyceride-associated variants with distinct body mass-versus body fat-related exposures as well as genotype-specific associations between alcohol consumption and liver stress at the *ADH1B* gene. Our catalog of vQTLs and GEIs is publicly available in an online portal.

## INTRODUCTION

Despite advances in identifying the genetic and environmental determinants of common complex diseases like cardiovascular disease and type 2 diabetes, the variability in the penetrance of genetic effects and the role played by environmental factors across populations are not fully understood. Part of this variability is due to gene-environment interactions (GEIs), in which genetic and non-genetic exposures synergistically affect disease-related traits. Understanding how exposures, including demographic, physiological, and lifestyle, modify genetic effects can spur new biological insights and therapies. Conversely, clarifying the ways in which one’s genetic background alters the effect of environmental exposures is a key step towards genome-guided precision medicine.

Comprehensive mapping of cardiometabolic GEIs across all genome-wide genetic variants and possible exposures carries practical, computational, and statistical challenges. Practically, it is difficult to collect and examine the thousands of exposures necessary for an “exposome-wide” approach, though recent software tools have made the process of high-dimensional phenotype processing substantially easier^1^.

Meanwhile, the massive number of GEI tests involved renders such an endeavor computationally infeasible and statistically underpowered, compounding known power limitations due to typically modest GEI effect sizes and low breadth and precision of exposure measurements^2^. Many screening procedures have been proposed to circumvent these computational and statistical limitations by reducing the genetic search space. These strategies prioritize specific sets of variants for GEI testing, such as those with main effects on the outcome^3^ or the exposure^4^.

GEIs may induce differences in the variance of continuous phenotypes across genotypes. Thus, tests for genetic markers associated with this variance, termed variance quantitative trait loci (vQTLs), represent an alternate strategy to identify loci harboring underlying GEIs for quantitative traits^5–11^. vQTLs can be identified in genome-wide scans analogous to those testing for phenotypic mean differences in typical genome-wide association studies (GWAS). Though direct GEI tests are more powerful when the environmental factor is measured accurately, vQTL tests may be advantageous when the relevant exposure is unknown or poorly measured, or when multiple exposures have aligned GEI effects at a locus. Such scenarios are quite common in practice due to the high dimensionality, dense correlation structure, and poor measurement of typical environmental exposures^12^. Recent studies in the UK Biobank (UKB) have demonstrated that a genome-wide vQTL discovery approach for anthropometric and lung function-related traits prioritizes variants enriched for GEI effects^10,11^. However, it is unclear whether vQTL effects are observed more broadly for cardiometabolic traits and to what degree the specific underlying GEI relationships can be uncovered using a more comprehensive array of environmental exposures.

Our objective was to identify genetic variants associated with the variance of a series of cardiometabolic blood biomarkers and to leverage these associations to efficiently detect underlying GEIs at an exposome-wide scale. We conducted a multi-ancestry, genome-wide vQTL scan to prioritize genetic loci across 20 biomarkers in UKB (N=350,016) in stage one, followed by an exposome-wide interaction study (EWIS), incorporating 2,380 exposures, to identify the specific underlying GEIs in stage two. We found 134 vQTLs, many of which were pleiotropic across biomarkers and largely overlapped with known GWAS loci. Using our EWIS approach, we then identified more than 800 specific GEIs with numerous correlated exposures underlying 54 of our vQTLs. Our study develops novel genetic maps of variance effects on a panel of cardiometabolic biomarkers, greatly increases the breadth of exposures tested for GEI, and introduces a publicly available catalog of vQTLs and GEIs that can inform precision medicine related to cardiometabolic health.

## RESULTS

### vQTLs are common and concentrated in known GWAS loci

The primary analysis consisted of two stages: first, the identification of vQTLs via genome-wide analysis using Levene’s test^13^, and second, the exploration of underlying GEIs at these loci across thousands of exposures (workflow described in Fig. 1). Each stage was conducted in each of four ancestry groups in the population-based UKB cohort (Supp. Table S1), though the European-ancestry subset was by far the largest, comprising 96% of the sample. Twenty cardiometabolic serum biomarkers were examined in this study, including lipids, lipoproteins, glycemic traits, liver enzymes, and kidney function markers (biomarkers listed in Supp. Table S2; preprocessed biomarker distributions shown in Supp. Fig. S1).

**Figure 1:**
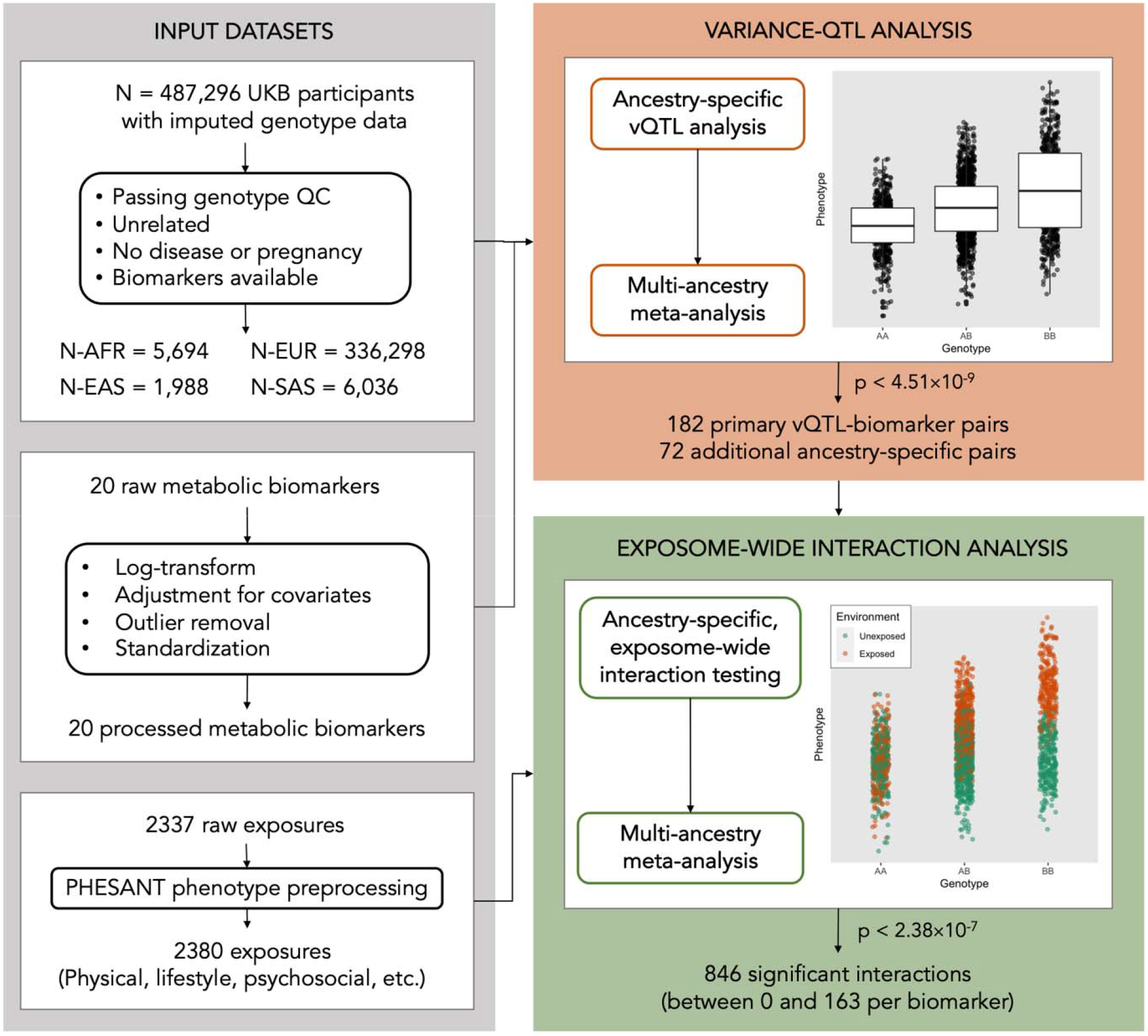
Analysis workflow.

In stage one, a study-wide Bonferroni significance threshold of *p* < 4.51×10^−9^ was established to correct for testing of 11.1 effective biomarkers (see Methods). The meta-analysis identified 182 vQTL-biomarker pairs at 134 independent loci after distance-based pruning (Fig. 2a, Supp. Table S3). While most vQTLs were biomarker-specific, a modest proportion were “pleiotropic” with respect to phenotypic variance, with five loci common to at least four biomarkers (Fig. 2b). The locus surrounding rs7259350 near the *APOE*/*APOC* cluster was the most pleiotropic, associating with the variance of nine biomarkers (including lipids [TC, LDL-C, HDL-C, TG], lipoproteins [ApoA, ApoB, LipA], liver enzymes [ALT], and hsCRP). This locus has strong main effects on the same biomarkers and is known to interact with lifestyle factors in determining cardiovascular disease risk^14^.

**Figure 2:**
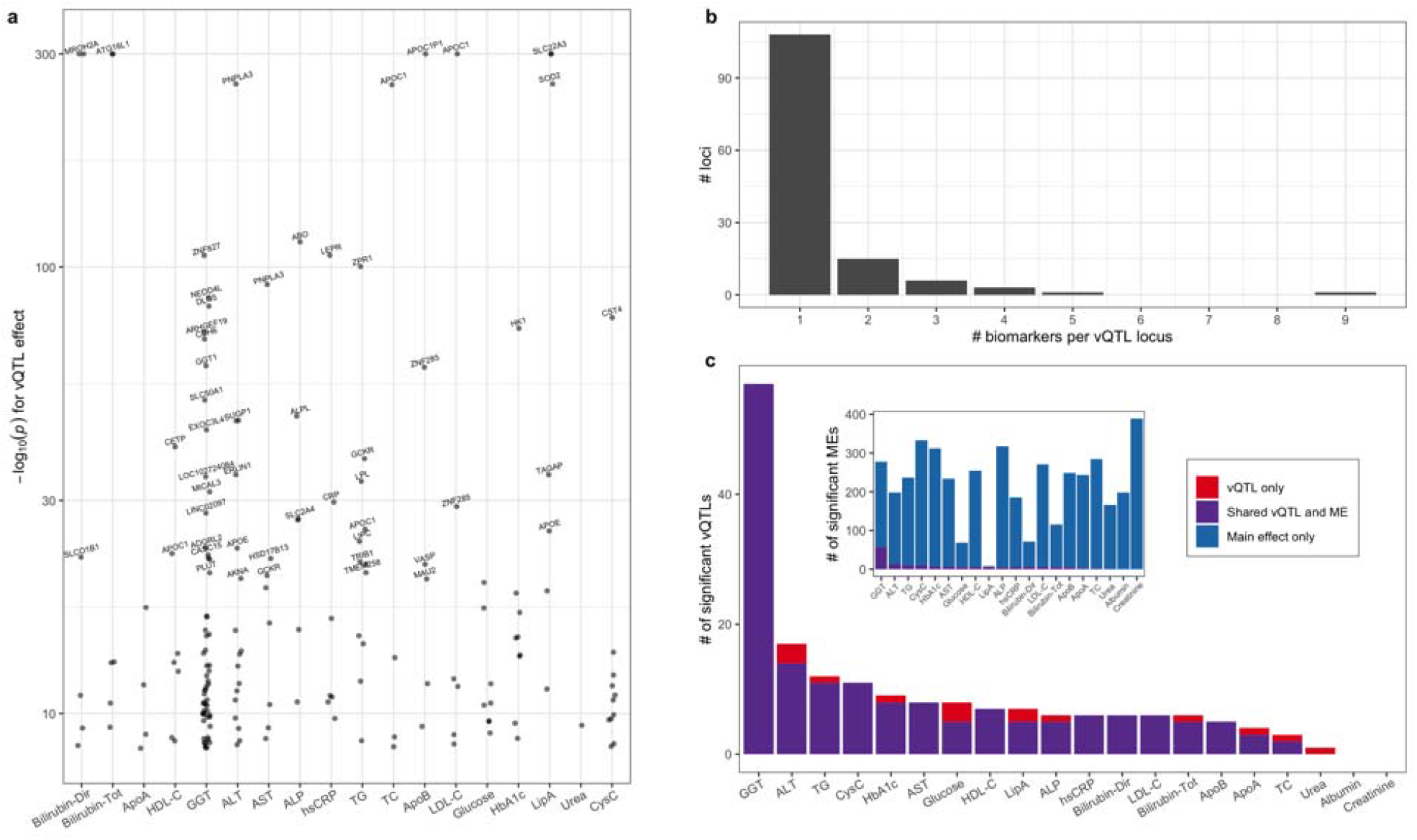
vQTLs identified across 20 cardiometabolic serum biomarkers. a) -log_10_*P*_vQTL_ is shown for all significant index variants for each biomarker. Labels correspond to the closest gene (shown for variants with *P*_vQTL_ < 10^−20^), highlighting some known GWAS loci. *P*-values are truncated at 10^−300^ for visualization purposes. b) Histogram displaying the number of biomarker associations for each vQTL locus. c) The number of significant vQTL loci is shown for each biomarker (inset: analogous plot for main effects). Colors denote three categories: vQTL loci not shared with an ME locus (red), vQTL loci shared with an ME locus (purple), and ME loci not shared with a vQTL locus (blue).

P-values from the vQTL meta-analysis tracked closely with those from the European subset (Supp. Fig. S2), as expected given the associated sample sizes. However, there were also 72 ancestry-specific vQTLs in one or more ancestry-specific analyses but not the meta-analysis (Supp. Table S4), 62 of which were found in non-European ancestry groups. While these ancestry-specific vQTLs may indicate the presence of heterogeneous variance effects across populations due to genetic ancestry differences or ethnic differences in environmental exposures, it is also possible that the non-European findings are the result of spurious associations given lower sample sizes, especially at lower minor allele frequencies^10^. Therefore, downstream analysis focuses on the meta-analysis vQTL findings only.

To understand these vQTLs in the context of genetic main effects (MEs), standard GWAS were also conducted for the same adjusted biomarker phenotypes. The resulting genetic MEs showed strong overlap with the identified vQTLs (Fig. 2c, Supp. Table S5). The majority of vQTLs had significant main effects: across all biomarkers, 91.6% of vQTLs were in ME loci. However, the converse was not true: only 3.7% of ME loci contained vQTLs. Thus, while this analysis did not discover many novel loci, it substantially narrowed the genomic search space and therefore multiple testing burden for downstream analysis as compared to starting with the set of loci from a standard mean-effect GWAS. Creatinine was particularly notable in this comparison, having the greatest number of MEs but no vQTLs – this could be explained by a true lack of underlying GEIs or gene-gene interactions, or by the limited power of vQTL approaches to detect more-complex interactions (e.g., involving multiple exposures in opposite directions). In contrast, Lipoprotein A was especially enriched for vQTLs, with 7 of its 8 MEs having vQTL associations. Beyond simple overlap of loci, we observed a quantitative relationship between vQTL and ME significances, which persisted when examining specific biomarkers whose raw values were either normally distributed (HbA1c) or non-normally distributed (GGT) (Supp. Fig. S3). This quantitative relationship confirms a similar result found for body mass index (BMI) in the UKB, and may be due to the fact that interactions are more likely to be present at loci with established biological connections to the phenotype of interest^11,15^.

We next conducted a genetic correlation (ρ_g_) analysis, using bivariate LD-score regression, to understand whether specific pairs of biomarkers were notably more or less similar in their genetic architecture when measured using MEs (standard) or vQTLs. We observed similar genetic correlations between biomarkers when using vQTL (ρ_g,vQTL_) and main-effect (ρ_g,ME_) summary statistics (Spearman correlation of 0.69 between ρ_g,vQTL_ and ρ_g,ME_ values across all non-identical biomarker pairs; Supp. Fig. S4). Genetic correlation magnitudes tended to be similar (mean |ρ_g_| of 0.16 across all non-identical biomarker pairs), while ρ_g,ME_ p-values were substantially lower. For nine biomarker pairs, the ρ_g,vQTL_ estimate was substantially higher (|ρ_g,vQTL_| - |ρ_g,ME_| > 0.2; such as between HbA1c and AST). These pairs may thus be more similar in their modifiable genetic effects (through GEIs, for example) than their fixed genetic effects.

We performed a replication analysis for each of the significant vQTLs for the 10 biomarkers available in the Women’s Genome Health Study (WGHS; N = 20,852 women of European ancestry)^16^. Of 61 significant vQTL-biomarker pairs for which replication was possible, nominal replication (*p* < 0.05) was seen for 27 (44%) in spite of the much smaller sample size (full set of vQTL replication results in Supp. Table S6). We found a strong correlation between discovery and replication significances (Spearman correlation of 0.49 between the p-values; Supp. Fig. S5a). The strongest vQTL associations in both the discovery and replication were with lipid-related biomarkers; for example, we find nominal replication for 5/6 LDL-C vQTLs, but only 1/9 HbA1c vQTLs.

### Exposome-wide interaction study reveals interactions underlying many vQTLs

In stage two, we conducted exposome-wide interaction tests for the 182 vQTL-biomarker pairs. We used the PHESANT program to produce 2,380 filtered and cleaned exposures, including physical, lifestyle, psychosocial, medical, and other types of traits (Supp. Table S7; see Methods). For each vQTL-biomarker pair, we tested for GEI effects involving the index variant and biomarker with each of 2380 exposure variables as potential effect modifiers, first stratified by ancestry and then meta-analyzed. Using a conservative multiple testing adjusted significance threshold (*p* < 0.05 / 182 / 1,156.2 effective exposures = 2.38×10^−7^) we identified 846 significant interaction effects at 54 of the 182 vQTL-biomarker pairs, altogether representing 34 loci (Fig. 3, Supp. Table S8). The 846 GEIs were unevenly distributed across biomarkers, with greater than one hundred for ALT, TG, and AST and none for albumin, creatinine, cystatin C, total bilirubin, and urea. Many of the participating exposures were from highly correlated categories such as anthropometric measures. This dense correlation structure means that many resulting interactions may reflect the same biological phenomenon, but also provides a unique opportunity to isolate the specific relevant exposure with greater precision.

**Figure 3:**
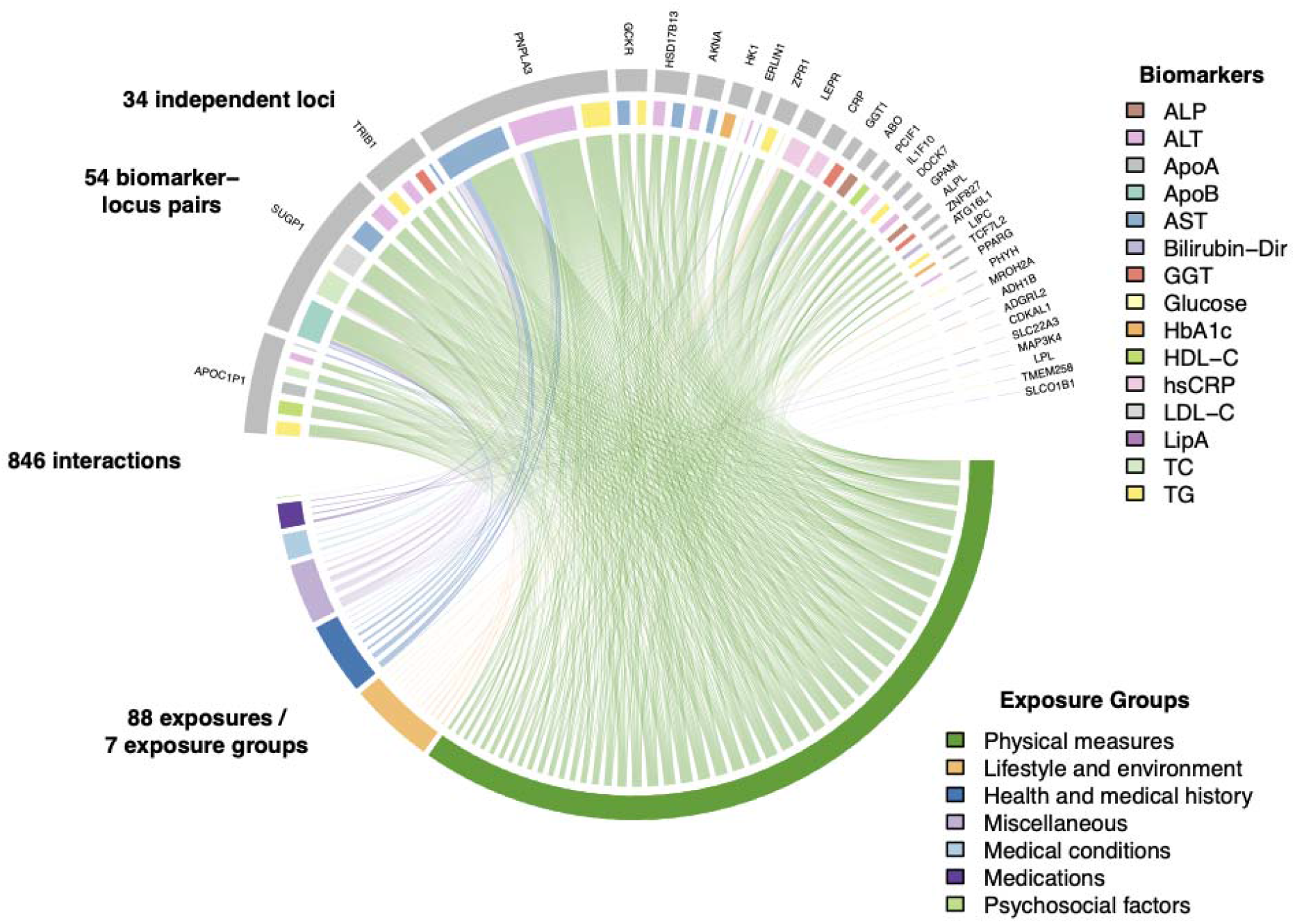
Chord diagram displays GEI links between vQTL-biomarker pairs (top of circle) and exposures (bottom of circle). Lines correspond to interactions that are Bonferroni significant (*p* < 2.38×10^−7^) for the associated variant, biomarker, and exposure. vQTL-biomarker pairs are colored according to biomarker and labeled with the nearest gene. Exposures are colored according to exposure categories.

Returning to the WGHS dataset, we chose to replicate specifically those interactions involving BMI as an exposure, since (1) many significant GEIs implicated BMI or another anthropometric trait (84%), (2) BMI is an objective and standard quantitative measurement that is easily compared across studies, and (3) BMI has strong biological links to the majority of biomarkers assessed in this study. Of 12 interactions involving BMI and one of the 10 available biomarkers in WGHS, nominal replication (*p* < 0.05) was seen for seven interactions (58%; Chi-square *p* = 2.31×10^−17^; Supp. Table S9). These included 3/5 (60%) BMI interactions for TG, 3/3 (100%) for hsCRP,1/1 (100%) for ApoB, and 0/1 for each of TC, HDL-C, and HbA1c. The replication of these interactions demonstrates their robustness across populations despite the substantially lower sample size in WGHS. As in the WGHS vQTL replication in stage one, we found that the strongest GEI signals by p-value from the discovery UKB cohort replicated in WGHS; in fact, the 7 BMI interactions that replicated were among the top 8 BMI interactions in UKB (among those with matching biomarkers in WGHS).

### vQTLs and interactions are robust in sensitivity analyses

To increase confidence in the catalog of vQTLs and interactions identified, we undertook a series of sensitivity analyses. First, we re-tested a subset of 174 vQTLs (those that were significant in the European-ancestry subset) using inverse-normal transformed (INT) biomarkers again in the European subset to confirm that the vQTLs were not artifacts of skewed biomarker distributions. While a substantial number of these relationships were attenuated, 47% remained significant at the Bonferroni level (94% at a nominal threshold of *p* < 0.05; Supp. Fig. S6).

Previous work has shown that naive two-stage, vQTL screening-based interaction testing procedures can have inflated type I error whenever the exposure tested in stage two is associated with the outcome, due to a correlation between test statistics for stage one and stage two^17^. In the context of a single exposure, one solution is straightforward: residualize the phenotype on the exposure prior to vQTL testing. However, this approach is not optimal when undertaking an unbiased EWIS with over 2,000 exposures: it is impractical to stably fit regression models with this dimensionality in the smaller non-European ancestry groups in this study (N < 6,100). Thus, we performed a sensitivity analysis based on the results of the primary analysis, residualizing each preprocessed biomarker on the much smaller set of 88 significant GEI exposures (separately in each ancestry) and re-performing genome-wide vQTL scans plus meta-analysis. In these exposure-adjusted analyses, 141 out of the original 182 vQTL-biomarker pairs (77%) remained significant (Supp. Fig. S7a), with only one completely diminished signal (p > 0.05). This result means that most of these vQTLs would have passed on to stage two for interaction testing even after explicit removal of this potential bias in the two-stage testing procedure. As additional support for the robustness of our interaction results to false positives, we did not observe substantial systematic inflation of interaction p-values across the entire set of EWIS tests (Supp. Fig. S7b).

### Investigation of anthropometric exposures for triglyceride GEIs highlights distinct biology

Many significant GEIs involved correlated anthropometric exposures, such as BMI, waist and hip circumference, and bioelectrical impedance metrics (which measure body fat and fat-free mass). Focusing on TG specifically as a biomarker with known complex relationships with obesity and cardiometabolic disease risk, we extracted GEI z-scores for the nine variants participating in significant interactions with an anthropometric trait (all of which are annotated to well-known TG-related genes). The nine variants showed distinct patterns of variation across exposures: two interacted more strongly with body fat measures (*FADS1-2* and *LIPC*), three interacted more strongly with body mass measures (*APOC1, ANGPTL3*, and *LPL*), and four were balanced across anthropometric exposures (*TRIB1, APOA5, GCKR*, and *PNPLA3*) (Fig. 4a). Given such highly correlated measurements, we also conducted a principal components analysis on the 33 relevant anthropometric exposures in the European subset, finding that top principal components appeared to represent body mass (PC1) and body fat (PC2; Supp. Fig. S8). These PCs reproduced the differential interactions with body mass and body fat (Fig. 4a).

**Figure 4:**
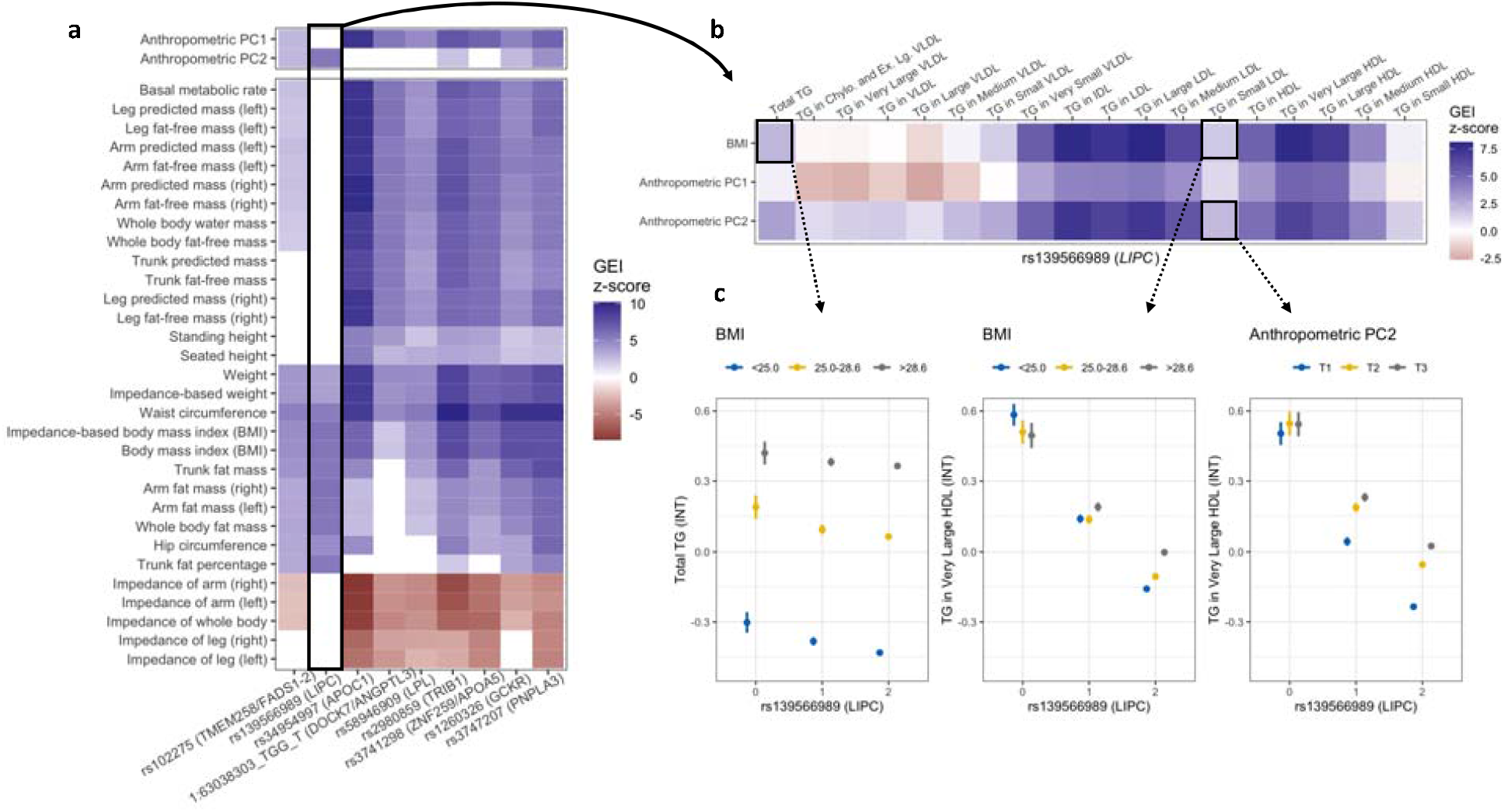
Exploration of anthropometric interactions influencing triglycerides. a) Heatmap shows interaction z-scores between nine genetic variants (*x*-axis) and 33 anthropometric exposures (*y*-axis). Colored panels pass a nominal significance threshold (*p* < 0.05). Variants are annotated with the closest gene, as well as a second likely causal gene based on manual annotation where appropriate. b) Heatmap shows interaction z-scores for a single variant (rs139566989 in *LIPC*), with varying TG lipid subfraction outcomes from nuclear magnetic resonance (*x*-axis) and three representative anthropometric exposures (*y*-axis). c) Three stratified plots showing means and 95% confidence intervals for inverse-normal transformed total TG or TG in very large HDL after stratification by rs139566989 and tertiles of the relevant exposure (labeled at the top of each plot).

The intronic indel rs139566989 in the hepatic lipase (*LIPC*) gene interacted almost exclusively with body fat-related measures. Hepatic lipase is a glycoprotein in the triacylglycerol lipase family that is important for the metabolism of lipoproteins including intermediate-density lipoprotein (IDL) and HDL^18^. The insertion (AF = 79% in the European subset) is associated with increased expression of *LIPC* (Genotype-Tissue Expression Portal browser) and decreased serum TG (*p* = 3.86×10^−59^ in our primary ME analysis). Its GEI effect with BMI represents a small but significant increase in the positive association between BMI and serum TG (*p* = 5.05×10^−25^). To further pinpoint relevant biological mechanisms, we further tested GEIs with BMI and the two anthropometric PCs for their effects on TG-containing lipid subfractions measured by nuclear magnetic resonance in the UKB (N∼90,000; Figure 4b). We found the strongest interactions (with both BMI and anthropometric PC2) for TG in IDL, large LDL, and large HDL (Fig. 4b). For example, the insertion increased the association between adiposity and TG in very large HDL: the bottom and top BMI tertiles had little TG difference in non-carriers, but a mean difference of almost 0.3 standard deviations in insertion homozygotes (Fig. 4c). Interactions between these three primary anthropometric exposures and all nine TG-related variants impacting TG subfractions can be found in Supp. Table S10.

A group of three variants (annotated to *APOC1, ANGPTL3*, and *LPL*) had a strong interaction with body mass PC1 on TG levels (all p ≤ 2.4×10^−6^) but none with body fat PC2. These three genes are involved in the production or regulation of lipoprotein lipase, which cleaves TG from circulating lipoproteins^18^. These body mass-specific interactions may reflect (1) biological processes in skeletal muscle rather than adipose, or (2) body mass-associated behavioral characteristics such as total caloric intake, which is likely better measured by body mass than from self-reported questionnaires.

### Alcohol intake interacts with a common ADH1B polymorphism to influence liver stress

One of our lead vQTLs with a clear underlying GEI was the combined effect of SNP rs1229984 and alcohol consumption on ALT, a biomarker of liver stress. SNP rs1229984 is a missense variant in the *ADH1B* gene that affects alcohol processing in the liver and is known to influence alcohol consumption^19^. This variant had a strong vQTL effect (*p* = 1.71×10^−13^) but did not have a significant main effect (*p* = 1.68×10^−4^) in the meta-analysis for ALT (Fig. 5a), highlighting the value in the vQTL screen for stage 1. In the following exposome-wide scan, the exposure most strongly interacting with rs1229984 to influence ALT was in fact self-reported “alcohol intake frequency” (*p* = 1.9×10^−12^; Fig. 5b). After stratifying self-reported alcohol intake into three bins within the European-ancestry group, we observed a substantial positive association between alcohol and ALT in homozygous major allele carriers, versus no such increase for minor allele carriers (Fig. 5c). The rs1229984-ALT vQTL was robust to pre-adjustment of the preprocessed ALT distribution for alcohol intake (*p* = 6.55×10^−13^ after adjustment), confirming that this original vQTL signal was not simply an artifact of the alcohol-ALT relationship. This interaction suggests that alcohol consumption may be of greater concern for potential liver damage in individuals homozygous for the major allele at rs1229984. We note that mean alcohol intake was much lower in T allele carriers, producing a decrease in intake variability that may contribute to the lack of alcohol-ALT relationship in these individuals.

**Figure 5:**
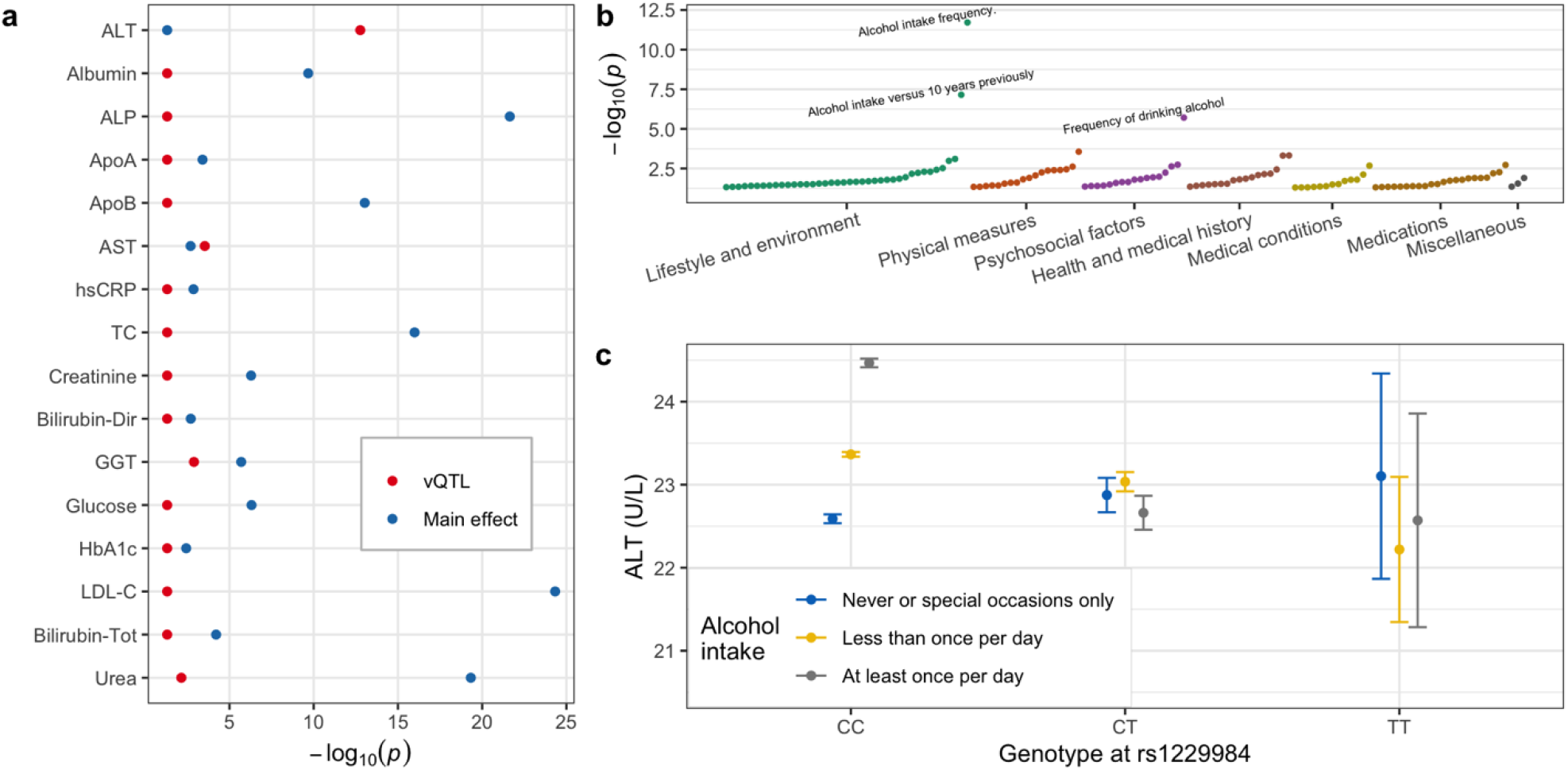
vQTL and GEI relationships for *ADH1B*, alcohol, and ALT. a) vQTL (red) and ME (blue) significance for rs1229984 is shown for each biomarker. Biomarkers with neither vQTL nor ME having *p* < 0.01 are not shown. b) EWIS results for rs1229984 impacting ALT are shown, with GEI significance plotted (y-axis) for each exposure having *p* < 0.05 (*x*-axis). c) Means and standard errors for ALT are plotted as a function of genotype at rs1229984 (*x*-axis) and self-reported alcohol intake (colors).

## DISCUSSION

We conducted a two-stage analysis to first identify vQTLs impacting cardiometabolic traits then conduct a systematic search for underlying GEIs. Using this strategy, we identified 134 loci associated with the variance of one or more of 20 cardiometabolic serum biomarkers, subsequently uncovering 846 GEIs impacting those biomarkers at 34 independent loci.

Results from our study largely align with previous results from Wang and colleagues, which utilized analogous methods in the same population (European-only subset of UKB)^10^. Examining anthropometric and lung function traits, they found a similar average number of vQTL relationships per effective phenotype (15 versus 16.4 in our study), indicating a similar level of polygenicity of vQTL effects. They found a somewhat smaller enrichment of vQTLs in ME loci (1.7% of ME loci contained vQTLs versus 4% in our study), suggesting that the genetic control of cardiometabolic biomarkers may be particularly susceptible to modulation by genetic background or environmental exposures compared to traits like height or forced vital capacity.

We observed that vQTLs were predominantly found in ME loci and that their strengths of association were generally correlated with the strengths of corresponding main effects. This observation has three important implications. First, it impacts the value of vQTLs as a screening tool. Screening that simply uses genetic main effects is a common and viable strategy^3^, but we note that (1) the presence of a vQTL further increases the likelihood of identifying an underlying interaction^11^, (2) in our analysis, the use of vQTLs reduced the genetic search space by more than an order of magnitude compared to genetic MEs, and (3) the alcohol-*ADH1B* interaction is an example of a strong interaction found via vQTLs that would not have been explored using only ME-based screening. Second, the correlation between vQTL and ME strengths could theoretically suggest that the vQTLs are statistical artifacts unrelated to underlying interactions. Non-normally distributed variables have a mean-variance relationship and thus some vQTL tests are susceptible to false positives in the presence of genetic MEs and can be sensitive to trait transformations^9,20^. However, previous studies have demonstrated reasonable robustness of Levene’s median-based test to non-normality^10^ and our sensitivity analyses demonstrate that inverse-normal transformed phenotypes, which have no mean-variance relationship, produce largely similar vQTL results. Third, it highlights the weight of prior probability in agnostic analyses: loci that have already demonstrated a role in biology (by being implicated in MEs) are more likely to be relevant in GEI searches as well.

Our two-stage approach required 4.3×10^5^ GEI tests, compared to the 4.8×10^11^ tests that would have been necessary to exhaustively explore all genetic variants and biomarkers. Beyond the clear practical and computational benefits of this reduction in the genetic search space, the statistical value in this two-stage analysis is dependent on both the power of Levene’s test and the extent to which it decreases the multiple testing penalty in the second stage (in this study, *p* < 2.38×10^−7^ compared to *p* < 3.90×10^−12^ without prioritization). While it is well-established that vQTL loci are more enriched than ME loci for underlying GEIs^10,11^, the power of Levene’s test may be affected by the strength of the underlying exposure-outcome relationship and may be compromised in cases where a complex set of underlying interactions exist without consistent directions of effect^6^. Given this fact, it is not surprising that many of our significant GEIs involved straightforward and high-impact exposures such as body size (e.g., BMI) and alcohol intake as compared to more complex traits like socioeconomic status indicators or dietary behaviors.

The PHESANT tool, which processes thousands of phenotypes in an automated way, was developed to facilitate phenome-wide association studies^1^, but here we leveraged it to create a library of exposures to be used in GEI tests. Many of its motivations apply equally in this context – non-normally distributed continuous variables and highly imbalanced binary variables can create instability and bias in GEI tests (as exposures) just as in standard genetic main effect tests (as outcomes). PHESANT enabled the high-throughput pre-processing of 2380 variables for our exposome-wide analysis, which would be impractical to prepare one-by-one. We note that the semi-automated nature of this process means that there may be more appropriate quality control and coding strategies for specific exposures of interest in follow-up studies.

Our approach identified 846 significant GEIs, many of which involved a set of highly correlated exposures (discussed further below). The large sample size of UKB provides the statistical power needed to identify interactions and has supported genome-wide GEI discovery in investigations of anthropometric and cardiometabolic phenotypes^21–24^. However, the scale of this population also means that spurious GEIs induced by even small statistical artifacts or biological confounders may reach statistical significance, as demonstrated by Tyrell and colleagues^25^. Thus, the catalog of GEIs described here should be treated as hypothesis-generating rather than confirmatory.

Many of the significant GEIs involved a set of highly correlated anthropometric exposures (e.g., BMI and waist circumference). These are not typically used as GEI exposures because they are not behavioral or environmental, but they can nonetheless can play a role in modifying genetic associations with cardiometabolic biomarkers and outcomes^26^. The dense correlation of these exposures reduces the number of effective discoveries, but this more-comprehensive coverage of the exposure space allowed us to tease apart underlying mechanisms. Focusing on TG as an outcome, we found that nine relevant variants tended to fall into three groups, interacting with anthropometric measures of either body mass, body fat, or both. We further validated this observation using representative principal component summary variables. For one variant (rs139566989 in *LIPC*), body fat interactions affected TG in specific lipoprotein subfractions such as IDL and very large HDL, consistent with the well-established role of hepatic lipase in lipoprotein remodeling (especially the IDL-to-LDL and large-to-small HDL conversions)^18^. Lipoprotein-related GEIs involving *LIPC* variants have been previously reported for relevant exposures such as dietary fat composition in weight loss trials^27,28^ and observational cohorts^29^. In general, our fine-grained exposure analysis provides insight into lipoprotein biology and highlights the importance of comprehensive and precise phenotyping.

A primary strength of this investigation is the comprehensiveness of cardiometabolic biomarkers and exposures examined. However, the pipeline relies on many self-reported exposure measurements; follow-up of specific interactions should employ more objective and precise methodologies (e.g., accelerometer-based physical activity measurements) as well as more comprehensive sensitivity analyses to refine interaction estimates. Additionally, we were only able to assess replication of vQTLs and GEIs for half of the biomarkers of interest. While the replication analysis provides confidence in the general robustness of our approach across populations, many signals from UKB, such as those for liver enzymes, bilirubin, and random glucose, were not able to be tested in WGHS. The WGHS also contains only women, which may particularly impact replication for sex-differentiated biomarkers such as HDL-C. Finally, while multiple ancestries were analyzed here as available, the meta-analysis results primarily reflect the effects in European individuals due to the major sample size discrepancy. Future work should incorporate more ancestry-balanced datasets, explore heterogeneity of interaction effects across ancestries, and pursue follow-up of some of the many ancestry-specific vQTLs and interactions described here.

The vQTLs and GEIs described here have been made publicly available in the AMP Common Metabolic Disease Knowledge Portal (https://hugeamp.org/research.html?pageid=UKB-vQTL-GxE). These catalogs can generate hypotheses for future research and enable lookups to better characterize findings from GWAS and other genetic studies. Our findings extend previous proofs-of-concept of the vQTL strategy to the realm of cardiometabolic biomarkers, establish a set of loci to be prioritized for further GEI analysis, and highlight specific GEIs that can inform cardiometabolic precision medicine strategies.

## METHODS

### UK Biobank genetic data

UKB is a large prospective cohort with both deep phenotyping and molecular data, including genome-wide genotyping, on over 500,000 individuals ages 40-69 living throughout the UK between 2006-2010^30^. Genotyping, imputation, and initial quality control on the genetic dataset have been described previously^31^. We removed individuals flagged for failing UKBiLEVE genotype quality control, heterozygosity or missingness outliers, individuals with putative sex chromosome aneuploidy, individuals with self-reported vs. genetically inferred sex mismatches, and individuals that had withdrawn consent by the time of analysis. Additionally, we subset to a group of unrelated samples by including only those that were used for genetic principal components analysis (PCA) during central genetic data preprocessing ^31^. Furthermore, only genetic variants with minor allele frequency (MAF) > 0.005 in the full sample were included in the present analysis (in addition to subsequent ancestry-specific MAF filters). For the vQTL and GWAS analyses, imputed genotypes within 0.1 of an integer value (0, 1, or 2) were converted to hard-calls using PLINK2 ^32^ and all other values were set to missing. Work was conducted on genetic data release version 3, with imputation to both Haplotype Reference Consortium and 1000 Genomes Project (1KGP), under UK Biobank application 27892. This work was conducted under a Not Human Subjects Research determination (NHSR-4298 at the Broad Institute of MIT and Harvard).

UKB samples were grouped into four primary ancestry groups: West African (AFR), East Asian (EAS), European (EUR) and South Asian (SAS). Using 1000 Genomes project (1KGP) phase 3 as the training dataset, probabilistic Gaussian mixture models were built to represent normally-distributed subpopulations within the overall population, assigning data points to the multivariate normal components that maximized the component posterior probability. Ten-fold cross-validation was used with different initialization states and clustering was evaluated using the adjusted Rand index score. The model with the highest adjusted Rand index score for each ancestry was used to cluster individuals in UKB into ancestry subgroups based on their 1KGP projected PCs and self-reported ethnicities (field 2100).

### Serum biomarkers

We focused on 20 serum biomarkers related to cardiovascular disease and metabolism, including but not limited to lipids, liver enzymes, glycemic parameters, and kidney function markers (see Supp. Table S2). Blood samples were collected at the baseline visit for the majority of participants, and specific biomarkers were measured using colorimetric, enzymatic, and immunoassays (details available at: https://biobank.ctsu.ox.ac.uk/crystal/crystal/docs/serum_biochemistry.pdf). Phenotype data processing and other downstream analyses were conducted using R version 3.6.0^33^ unless described otherwise. Biomarker values were pre-processed separately within each ancestry group, and followed a modified version of the procedure described by Sinnott-Armstrong and colleagues; see original manuscript for detailed description and online code^34^. We additionally removed individuals with diabetes, coronary heart disease, cirrhosis, end-stage renal disease, cancer diagnosis within one year prior to their assessment center visit, or who were pregnant within one year of the assessment center visit. Briefly, log-transformed biomarkers were adjusted for sex, age, self-identified ethnicity, fasting time, dilution factor, assessment center, genotyping batch, genetic principal components, time of sampling, month and day of assessment, and a series of interactions between covariates. Of note, cholesterol, LDL-C, and Apolipoprotein B were also adjusted for statin use using methods described previously^34^.

Following residualization, outliers (residuals greater than 5 standard deviations from the mean) were set to missing. Finally, residuals were z-score normalized to mean zero and variance one.

Due to the substantial correlation between the 20 cardiometabolic biomarkers, a smaller number of “effective” biomarkers was calculated to inform multiple hypothesis testing correction. Using a previously-described approach^35^, preprocessed biomarkers were collected into a single dataset across ancestries and principal components analysis was performed (*prcomp* function with standardized variables). The number of effective biomarkers was then calculated from the principal component variances λ (equal to the eigenvalues of the biomarker covariance matrix) as 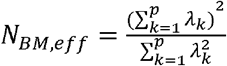.

### Variance-QTL analysis

vQTL analysis was performed using the vQTL module from the OSCA suite ^36^. Differential variance tests used the median-based Levene’s test, which is equivalent to a one-way analysis of variance (ANOVA) for absolute deviations from the median biomarker value. The test statistic, which is described in detail elsewhere ^10^, is:

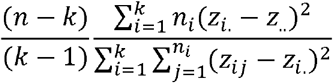

where *i* is the group indicator (one of three genotypes), *n*_*i*_ is the sample size for the *i*^*th*^ genotype, *z*_*ij*_ is the absolute difference between the phenotype value in sample *j* from genotype *i* and the median value in genotype *i, z*_*i*._ is the average *z* value in genotype *i*, and *z*_.._ is the average z value across all samples. Under the null hypothesis of equal variances, this statistic follows an *F* distribution with *k-1* and *n -k* degrees of freedom.

Genome-wide vQTL analysis was performed separately for each of the 20 biomarkers (Supp. Table S2) in each of the four ancestry groups. Multi-ancestry meta-analysis was then performed using METAL (2011-03-25 version)^37^. The inverse variance-weighted, fixed-effects meta-analysis strategy (based on effect sizes and standard errors rather than p-values and sample sizes) was used to incorporate effect directions. While Levene’s test does not natively produce effect sizes, they are derived by the OSCA tool based on a method described by Zhu and colleagues^38^ and previously implemented for vQTLs^10^. An effect direction is determined by regression of absolute deviations from the median on additively-coded genotypes, then this sign is combined with the Levene’s test p-value to derive an effect size and standard error through back-transformation: 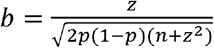 and 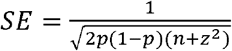 where *b* is the vQTL effect size, *z* is a z-statistic derived from the *p*-value and effect direction, *p* is the minor allele frequency of the variant, and *n* is the sample size.

For each genome-wide vQTL scan (20 biomarkers × 4 ancestries), we performed an analogous standard GWAS testing for genetic effects on the mean of the phenotype using the *--glm* command in PLINK2^32^. These GWAS used the same adjusted phenotypes with no additional covariates, followed by a fixed-effects, standard error-based, multi-ancestry meta-analysis approach as described above.

Genome-wide summary statistics were pruned by removing variants within 500kb of each index variant. This distance-based procedure was chosen to be reasonably conservative and avoid the need for separate LD matrices per ancestry. To identify overlapping variants across biomarkers, ancestries, and analysis types (vQTL vs. ME), relevant pruned summary statistics were combined and a second clumping procedure (similarly defined based on 1MB windows) was performed. For example, to identify overlapping vQTL loci across ancestries for ALT, all ancestry-specific, pruned summary statistic matrices for ALT (four in total) were stacked and then subject to a similar iterative clumping procedure as was used for the initial pruning.

### Preparation of exposure variables

In order to conduct the GEI analysis in an exposome-wide manner, we needed to collect and clean a large set of relevant exposure variables. For this purpose, we used a program for automated phenotype pre-processing initially developed for phenome-wide association studies (PHEnome Scan ANalysis Tool, or PHESANT ^1^) and later expanded (https://github.com/astheeggeggs/PHESANT). This PHESANT procedure relies on files describing variable input types (continuous, integer, categorical single [one choice], and categorical multiple [multiple choice]) and data coding schemes. Based on this input, it then conducts automated pre-processing of the phenotypic dataset, including removal of variables with excessive missingness or insufficient heterogeneity, conversion of categorical variables to ordinal and binary variables, and inverse-rank normal transformation of continuous variables.

As originally applied, the PHESANT pipeline processed a comprehensive set of phenotypes appropriate for phenome-wide association studies. We modified the set of included variables to better reflect the space of traits and exposures that may participate in GEIs. For example, sex does not make sense to use as an outcome, but may be an important characteristic that modifies genetic associations with blood biomarker levels. Specifically, in comparison to the PHESANT pipeline referenced above, we additionally included sex, age, and assessment center, and 24-hour recall-based dietary assessments and excluded blood-based assays, hospital records, and cancer and death registers. After running PHESANT on the full UK Biobank dataset, the output dataset contained 2,380 variables for analysis. This corresponded to 1,156.1 effective exposures tested based on the same procedure used above for determining the number of effective biomarkers. These exposures included physical (e.g., BMI), lifestyle (e.g., dietary behaviors), psychosocial (e.g., neuroticism score), medical (e.g., statin use), and other types of traits. Based on the data dictionary available from UKB and additional manual curation, phenotypes were placed into one of eight exposure categories. Exposure group summary counts and variable examples are available in Supp. Table S7.

### Genome-wide interaction studies

For each combination of vQTL-biomarker pair (from Stage I) and exposure (from PHESANT), a GEI test was performed using the following simple model:

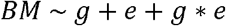

where *BM* represents the preprocessed biomarker, *g* represents the imputed genotype dosage at the vQTL index variant, and *e* represents the relevant exposure. For the primary analysis, no additional covariates were used given the prior residualization on basic covariates (age, sex, and genetic principal components). Interaction analysis was performed with GEM v1.3^39^ using robust standard errors. As with the vQTL and GWAS analyses, these interaction tests were performed separately in each ancestry group, followed by the inverse variance-weighted, fixed-effect meta-analysis^37^. Principal components analysis using prcomp in R was conducted on a set of anthropometric exposures in individuals of European ancestry in UK Biobank to include as additional body size-related exposures in GEI testing.

### Anthropometric exposure and lipid subfraction analysis

Anthropometric exposures were defined as those with a “Level 3 Category” of “Anthropometry” in the UKB data dictionary. Initially, GEI summary statistics from the EWIS were collected for this set of exposures with TG as the outcome and including nine genetic variants passing the EWIS significance threshold for at least one of these 31 exposures. These nine variants were manually annotated to likely causal genes based on known lipid biology. To create a set of anthropometric summary variables for GEI analysis, PCA was performed on this set of exposures in the European ancestry subset. The first two principal components were determined to represent body mass and body fat based on inspecting of trait loadings. These two components were extracted and used for GEI testing.

Triglyceride concentrations in various lipoprotein subfractions, based on nuclear magnetic resonance (NMR) spectroscopy metabolomics data, were retrieved for approximately 90,000 European ancestry individuals (differing numbers per metabolite based on missingness). Quality control was performed using measures provided by the UKBB, removing values lower than the limit of detection and samples with likely contamination or degradation issues. Metabolite measurements were inverse-normal transformed prior to modeling. GEI testing was performed for each anthropometric exposure-TG subfraction pair (as exposure and outcome, respectively), with additional adjustment for age, sex, 10 genetic PCs, NMR batch, and spectrometer.

### WGHS replication analysis

The WGHS cohort was used to replicate vQTLs and BMI GEIs. WGHS is a prospective US-based cohort of healthy adult females 45 years or older^40^. The biomarkers available in WGHS included ApoA, ApoB, hsCRP, TC, creatinine, HbA1c, HDL-C, LDL-C, LipA, and TG, and were pre-processed and analyzed similarly to biomarkers in UKB (limiting to European ancestry individuals, log transforming, removing individuals with diabetes or on blood pressure medication, adjusting for covariates, and testing vQTL hard-call genotype associations in OSCA; N=20,852). The same pre-processed biomarkers were used for replicating the BMI GEIs, adjusting for age and genetic PCs.

## Data Availability

UK Biobank data can be accessed by application directly at http://www.ukbiobank.ac.uk. Results from this analysis can be explored and downloaded at: https://hugeamp.org/research.html?pageid=UKB-vQTL-GxE.

## AUTHOR CONTRIBUTIONS

KEW, AKM, and JBC designed the experiment. KEW and JBC conducted the primary analysis. TDM conducted the UK Biobank metabolomic data analysis. HC and AKM contributed to the design and interpretation of the two-stage analysis approach. FG and DIC contributed to the replication analysis. JCF provided UK Biobank data access. JCF and MSU provided clinical interpretation of findings. KEW, DKJ, and JBC contributed to the storage and visualization of these results in an online catalog. MSU, DIC, AKM, and JBC supervised the analysis and the provided critical revision of the manuscript. KEW and JBC wrote the manuscript. All authors reviewed and approved the final manuscript.

## ACKNOWLEDGEMENTS

The authors would like to thank Jordi Merino of Massachusetts General Hospital and Chirag J. Patel of Harvard Medical School for helpful comments on the analysis.

KEW was supported by NIDDK Fellowship 2T32DK007028-46. HC and AKM are supported by NHLBI R01 HL145025. JCF is supported by NHLBI K24 HL157960. MSU is supported in part by NIDDK K23DK114551. JBC is supported by NIDDK Pathway to Independence Award (K99DK127196).

The WGHS is supported by the National Heart, Lung, and Blood Institute (HL043851 and HL080467) and the National Cancer Institute (CA047988 and UM1CA182913), with funding for genotyping provided by Amgen.

The Genotype-Tissue Expression (GTEx) Project was supported by the Common Fund of the Office of the Director of the National Institutes of Health, and by NCI, NHGRI, NHLBI, NIDA, NIMH, and NINDS. The data used for the analyses described in this manuscript were obtained from the GTEx Portal on 10/18/2021.

While KEW and AKM are employees of Mass General Brigham, this work was not conducted in their capacity as Mass General Brigham employees.

