## Supplementary Materials for "Variance-quantitative trait loci enable systematic discovery of gene-environment interactions for cardiometabolic serum biomarkers"

**SUPPLEMENTARY FIGURES**

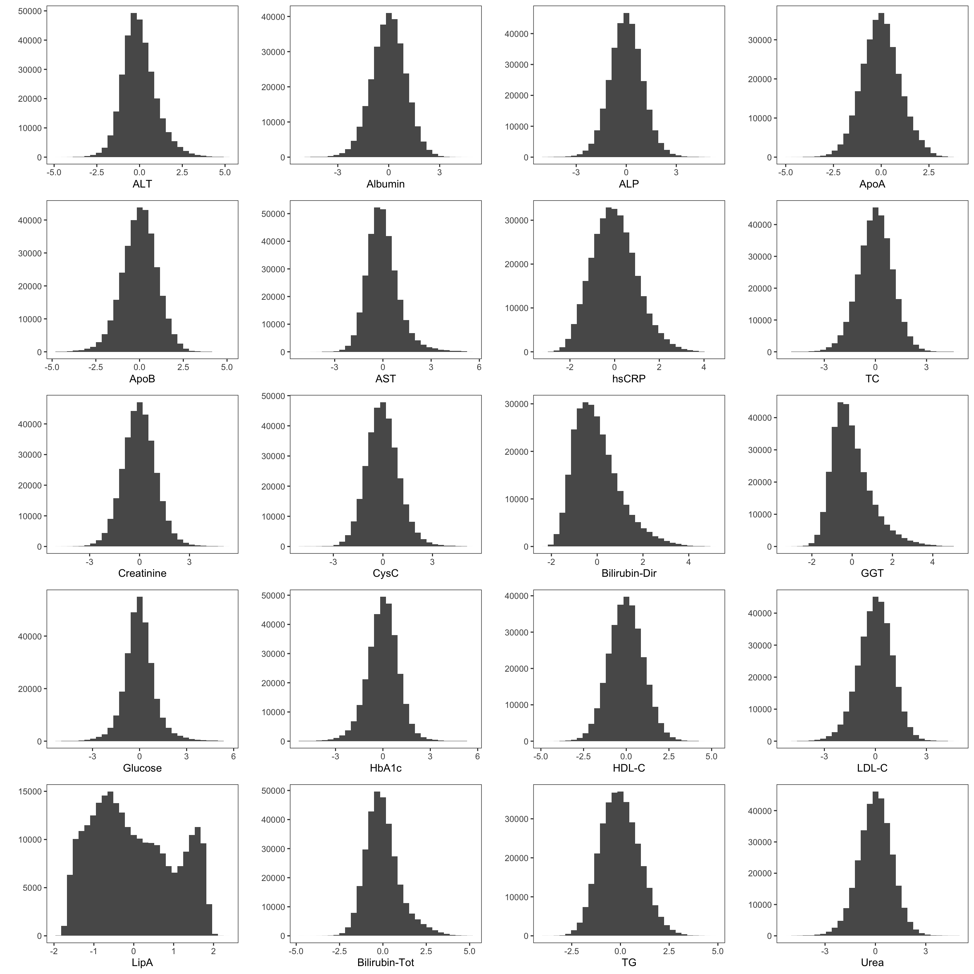

**Supplementary Figure S1:** Preprocessed cardiometabolic biomarker distributions for the European-ancestry population subset. Histograms display the counts of biomarker values used as input to the vQTL analysis for each of the 20 biomarkers. Preprocessing consisted of medication adjustment (where appropriate), log-transformation, adjustment for covariates, winsorization, and standardization to mean zero and standard deviation one.

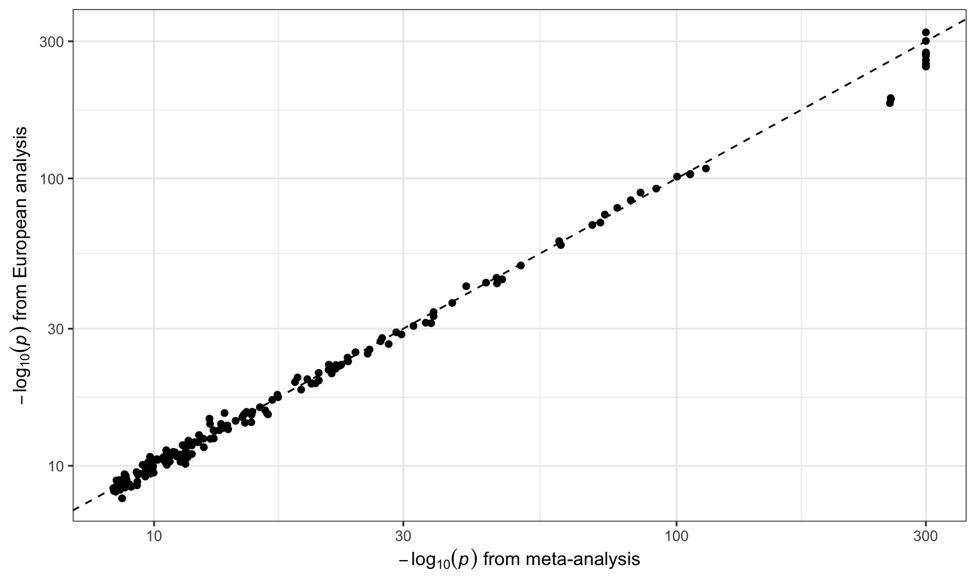

**Supplementary Figure S2:** Comparison of -log(p-value) between the vQTL meta-analysis and European-only analysis for each of the significant vQTLs from the meta-analysis. Dashed line denotes *y* = *x*. *P*-values are truncated at 10^-300^ for visualization purposes.

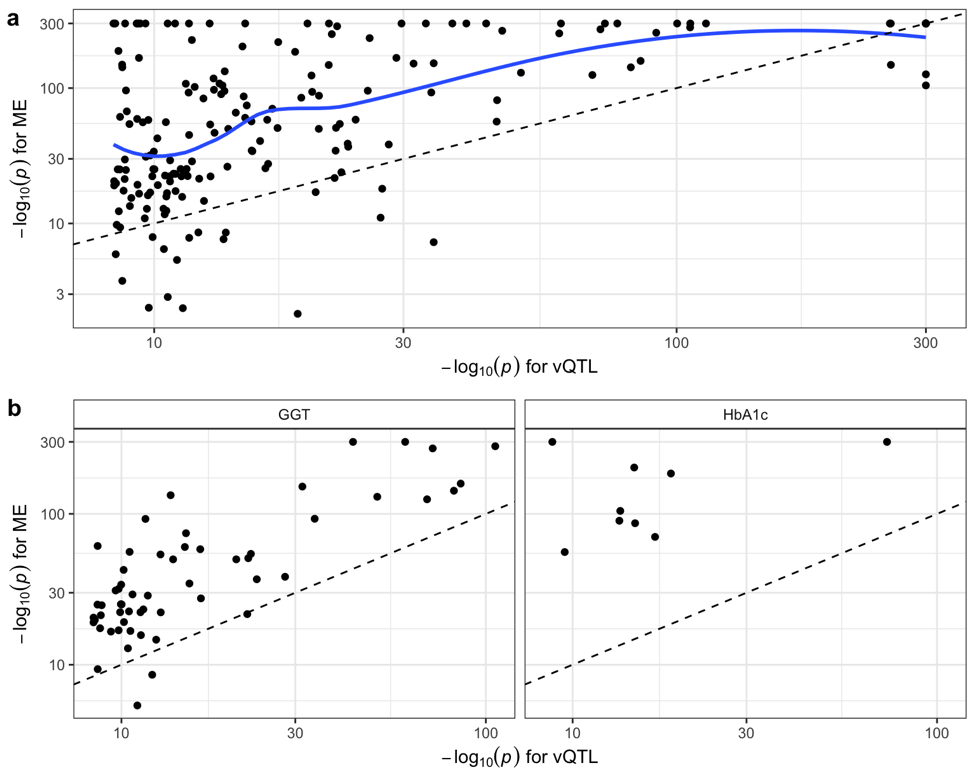

**Supplementary Figure S3:** Comparison of vQTL and main effect strengths across all biomarkers. *x*-axis and *y*-axis denote the significance of the associated vQTL and ME test, respectively, for each of the significant vQTLs. Dashed line denotes *y* = *x*. Plots are shown for (a) all vQTLs, and (b) vQTLs for GGT (a representative biomarker whose raw values are substantially skewed) and HbA1c (a representative biomarker whose raw values are approximately normally-distributed). Blue line in (a) is a LOESS curve of best fit. *P*-values are truncated 10^-300^ for visualization purposes.

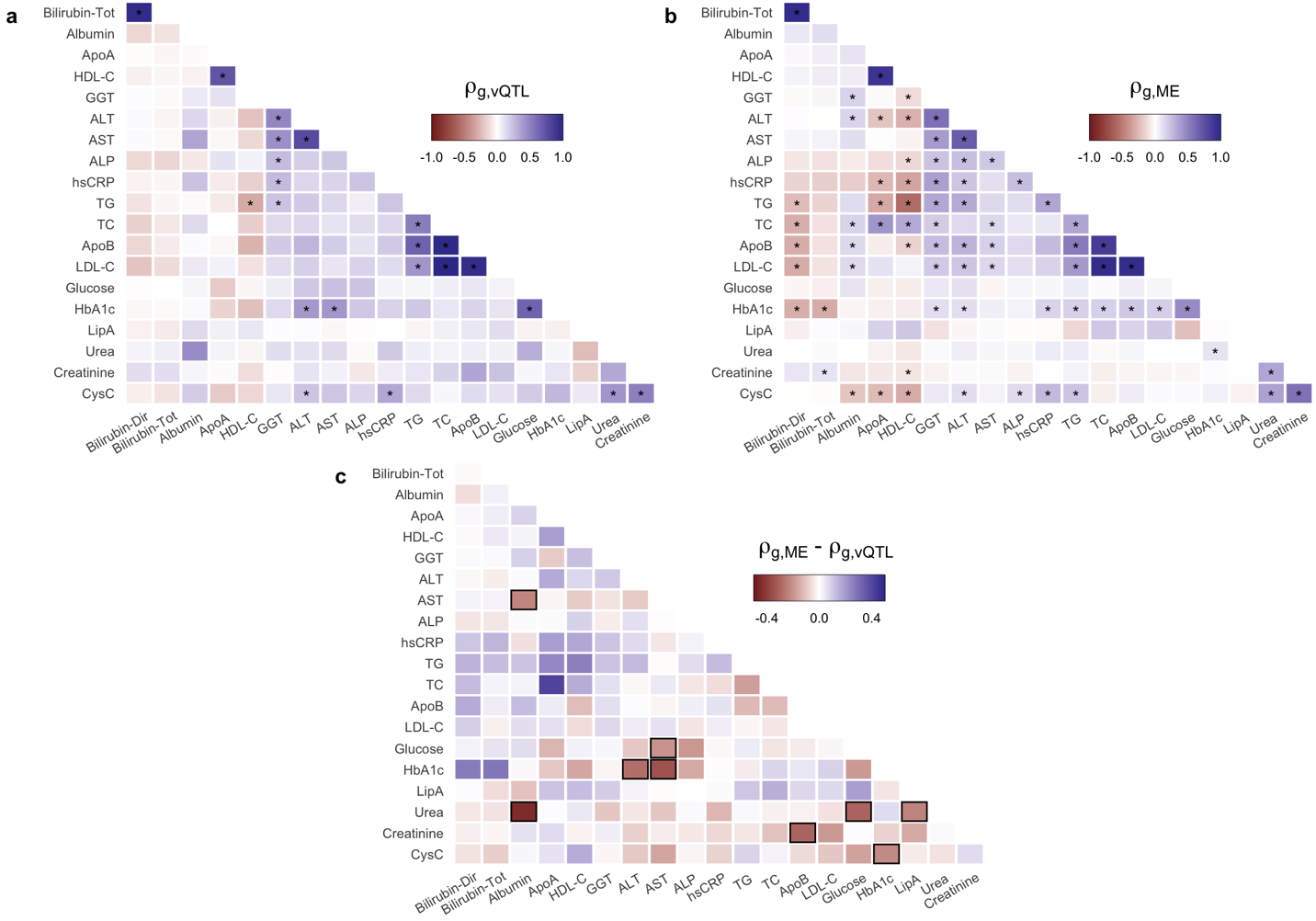

**Supplementary Figure S4:** Genetic correlations calculated using LDSC based on either vQTL summary statistics (a) or ME summary statistics (b). Heatmap colors denote the genetic correlation estimate, and stars denote significant genetic correlations after Bonferroni correction for the number of biomarker-biomarker comparisons (*p* < 0.05 / 190 = 2.6×10^-4^). Panel (c) shows the difference between the absolute value of the ME-based and vQTL-based genetic correlations, such that positive values represent stronger magnitudes of ME-based genetic correlation. Biomarker pairs for which the vQTL-based genetic correlation magnitude is greater by at least 0.2 are highlighted.

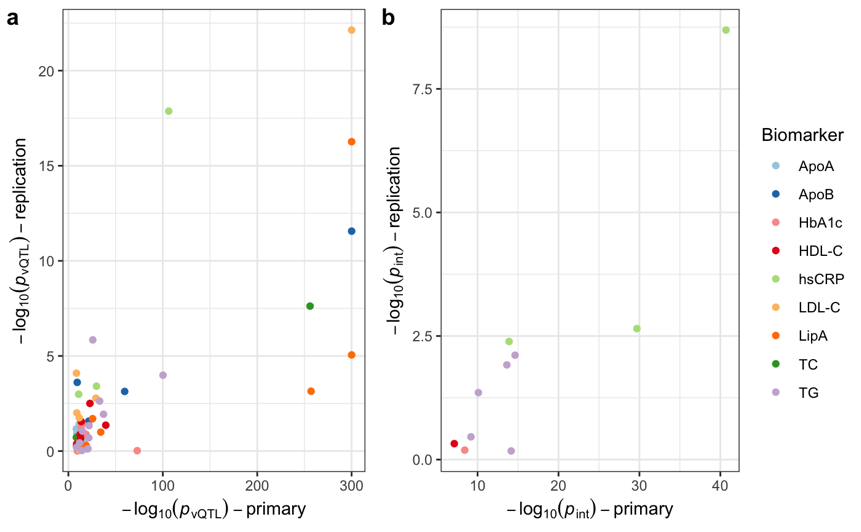

**Supplementary Figure S5:** Comparison of primary and replication p-values for vQTLs and GEIs. (a) Comparison of p-values from the primary analysis (*x*-axis) and the replication analysis in WGHS (*y*-axis). Points represent vQTL-biomarker pairs, and colors correspond to the relevant biomarker. P-values were truncated at 1×10^-300^ for plotting purposes. (b) As in (a), but instead comparing p-values for interaction with BMI as the exposure.

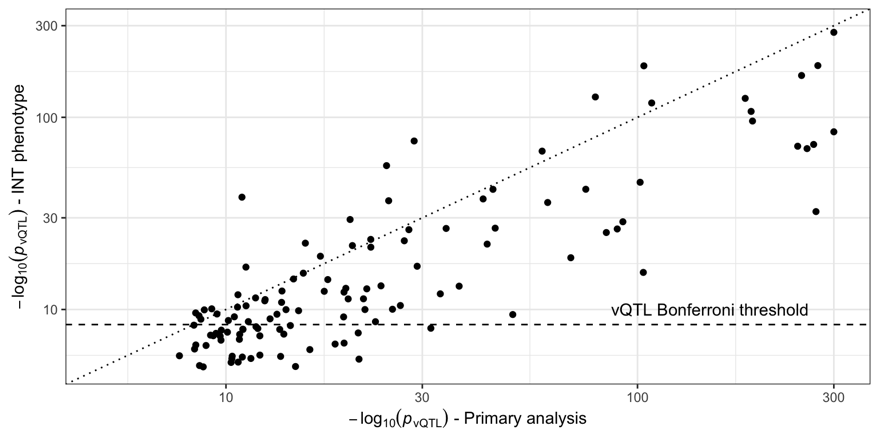

**Supplementary Figure S6:** Sensitivity analysis for vQTLs. Comparison of vQTL p-values from Levene’s test using preprocessed biomarkers from the primary analysis (*x*-axis) vs. additionally inverse-normal transformed versions of the same biomarkers (*y*-axis) for all significant vQTLs from the primary analysis. Values shown here are from analyses in the European ancestry group.

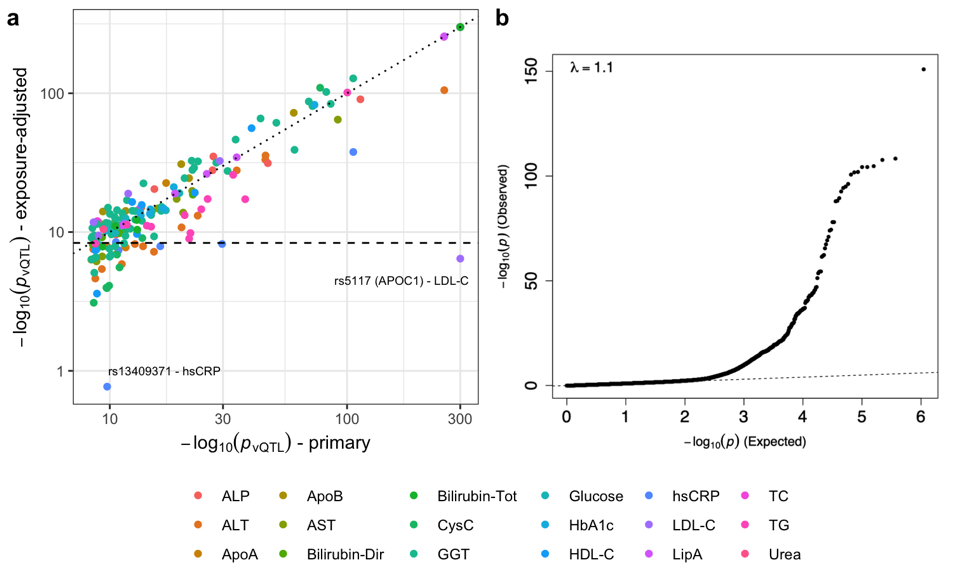

**Supplementary Figure S7:** Assessment of bias in exposome-wide interaction testing. (a) Negative log-transformed vQTL p-values are compared between the primary analysis and a sensitivity analysis using biomarker phenotypes further adjusted for the 88 significant exposures from the primary analysis. (b) Overall inflation of p-values from exposome-wide interaction testing. The quantile-quantile plot compares observed p-values (negative log-transformed; *y*-axis) to those expected based on a uniform distribution of p-values (*x*-axis). The lambda value denotes the genomic inflation value (ratio of observed to expected median Chi-square statistics).

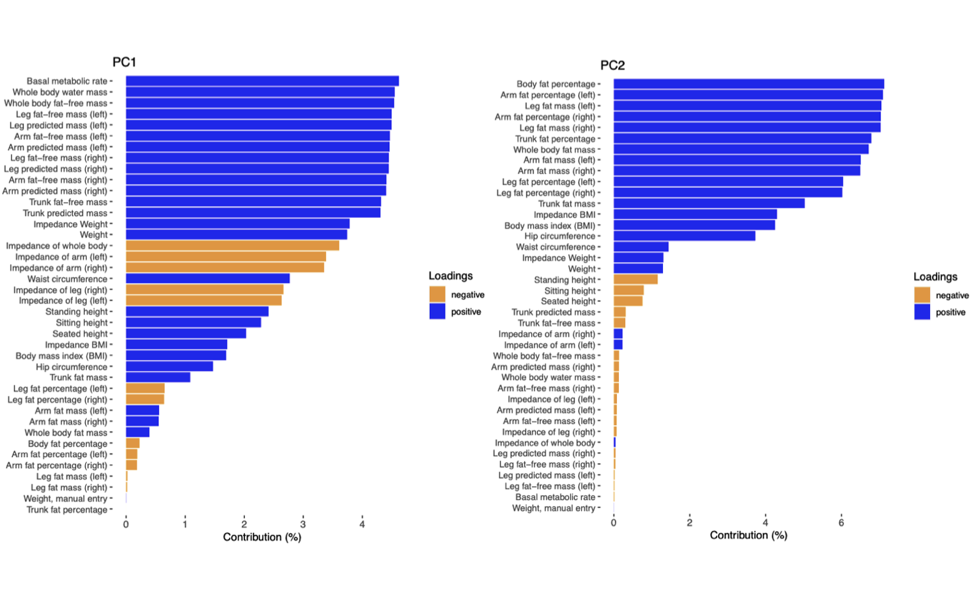

**Supplementary Figure S8:** Description of anthropometric principal components. Principal components analysis was run on anthropometric exposures, and the resulting top principal components explained 53.4% and 32.4% of the total variance. PC loading plots display the contribution (squared correlations) of individual traits to PC1 (left; primarily body mass-related) and PC2 (right; primarily body fat-related).

**SUPPLEMENTARY TABLES** (see accompanying Excel file)

**Supplementary Table S1:** Sample characteristics and biomarker levels by population.

Note: sample sizes correspond to individuals without cardiometabolic disease or recent cancer, not pregnant, and having biomarker and genetic data available, but these numbers differ across biomarkers for vQTL and EWIS analyses due to biomarker-specific missingness and outliers. Continuous values are presented as: mean (standard deviation).

**Supplementary Table S2:** Cardiometabolic biomarker details.

**Supplementary Table S3:** Significant vQTLs from multi-ancestry meta-analysis across all biomarkers.

**Supplementary Table S4:** Significant ancestry-specific vQTLs (loci not reaching significance in the meta-analysis) across all biomarkers.

**Supplementary Table S5:** vQTL and ME locus counts and overlap.

**Supplementary Table S6:** Replication of vQTL effects in WGHS.

**Supplementary Table S7:** Overview of exposures as derived from PHESANT.

**Supplementary Table S8:** Significant interactions from the exposome-wide interaction study.

**Supplementary Table S9:** Replication of BMI interactions in WGHS.

**Supplementary Table S10:** All interaction testing results for anthropometric exposures impacting TG subfractions.
